# Anticipating the hospital burden of future COVID-19 epidemic waves

**DOI:** 10.1101/2021.08.19.21262280

**Authors:** Corentin Boennec, Samuel Alizon, Mircea T Sofonea

**Author notes:** These authors contributed equally to this work (Samuel Alizon, Mircea T Sofonea).

## Abstract

Forecasting SARS-CoV-2 epidemic trends with confidence more than a few weeks ahead is almost impossible as these entirely depend on political decisions. We address this problem by investigating the consequences for the health system of an epidemic wave of a given size. This approach yields semi-quantitative results that depend on the proportion of the population already infected and vaccinated. We introduce the COVimpact software, which allows users to visualise estimated numbers of ICU admissions, deaths, and infections stratified by age class at the French departmental, regional, or national level caused by the wave. We illustrate the usefulness of our approach by showing that for France, even with a 95% vaccination coverage, the current vaccine efficiency against the delta variant would make a large epidemic wave infecting 25% of the population difficult to sustain for the current hospital bed occupancy capacity. Overall, using the final epidemic wave size and ignoring detailed epidemiological dynamics yields valuable and practical insights to optimise public health response to epidemics.

## 1 Introduction

In France, as in other countries, detailed mechanistic models have been developed that can capture COVID-19 epidemic dynamics, especially hospital admission and bed occupancy data [8, 10]. The problem is that these models are extremely unreliable to make forecasting more than 3 to 4 weeks ahead because epidemic dynamics rely on political decisions. For example, let us consider a country with an intermediate SARS-CoV-2 infection prevalence that decides to switch public health strategy the next week. It is obvious that the number of hospital admissions a month later will be strikingly different if the strategy consists in alleviating control over transmission chains (e.g. as some regions in Brazil in 2020), or in maintaining incidence to a high plateau (as France in spring 2021), or in applying ‘zero COVID’ strategy (as in countries such as New-Zealand, Australia, or Vietnam). In a way, COVID-19 epidemiological forecasting more than a month ahead largely consists in anticipating political decisions. On top of this, additional uncertainty originates from social (e.g. school holidays), environmental (e.g. humidity), and biological (e.g. virus evolution) factors.

We address this issue by ignoring detailed temporal epidemiological dynamics and summarising these into a single metric namely the final epidemic size, i.e. the proportion of the population that will be infected during the epidemic wave. This allows us to estimate the resulting number of casualties and of intensive care unit (ICU) admissions, one of the most crucial indicators for health care systems, caused by a future epidemic wave.

Several hurdles need to be overcome that are further discussed in the Methods and in Supplementary Text S1. First, we need to stratify the total number of cases in the epidemic wave by age the SARS-CoV-2 infection fatality ratio is strongly age-dependent [8]. Second, and also for each age, we need to estimate which proportion of the population that has already been infected by the virus and has developed immunity. Third, we need to set the proportion of the population has been vaccinated, and by which type of vaccine. Again, this variable should be stratified by age. To help pubic health field workers, we also introduce a dedicated software COVimpact, which performs these estimations at the French department, regional, and national level. In spite of the lack of temporal dynamics (we do not know whether the total number of admissions will be spread over one or 4 months), the semi-quantitative results we obtain provide valuable insights on the long term (more than a month ahead). As we illustrate in the case of France, where our results indicate that vaccination alone is unlikely to be sufficient in the near future, this approach can be instrumental in planning and optimisation local hospital response to future COVID-19 epidemic waves.

### 2 Methods

Our goal is to estimate the total number of ICU admissions and deaths caused by an epidemic wave of a given size, i.e. associated with the total number of people infected. A first difficulty is that this variable, as most of the ones used in our approach, needs to be stratified by the age of the individuals (*a*) and their geographical location (department, region, or country). Two other layers of stratification are natural immunity (i.e. having recovered from a previous infection) and vaccine immunity. If *N* vaccines are being implemented in the population, and assuming an individual is only vaccinated by a single type of vaccine, this leads to 2*N* + 2 host categories for each age.

The number of hosts of age *a* that have been infected in the past is calculated by combining a previous mathematical model from the team [10] and detailed data analysed by Hozé et al. [4]. Note that some hosts can be reinfected (approximately 16% [2]), which means that the number of hosts with natural immunity is lower than the number of past infections.

The proportion of hosts of age *a* vaccinated by vaccine *i* is set by the user. The vaccine efficacy, i.e. the protection against severe infection forms, is also set by the user. Note that our model formalism does not require any assumption regarding vaccine protection against reinfection or against transmission. We assume that vaccination is not targeted towards hosts without natural immunity. For hosts with both natural and vaccine immunity, we assume that the level of protection conferred against severe infection forms is the highest of the two protections.

Finally, we assume that all hosts are exposed in the same way to the infection, i.e. there is no effect of age, or of natural and vaccine immunity.

Regarding the link between the epidemic size, estimating the proportion of hosts infected by an uncontrolled epidemic wave (*p*\*) is an old problem in epidemiology and it is known to be tightly linked with the basic reproduction number (*R*_0_), the expected number of secondary infections caused by an infected host in an otherwise fully susceptible population. Mathematically, for a simple Susceptible-Infected-Recovered model, these variables are linked through a simple formula which in this work [5]:

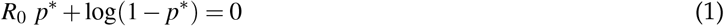

Additional details about the methods are in Supplementary Text S1 and the scripts used can be accessed at https://gitlab.in2p3.fr/ete/residual_epidemic.

## 3 Results

We first present the dedicated software for the French epidemic, which can be accessed at https://cloudapps. france-bioinformatique.fr/covimpact/, before focusing on a scenario for the French epidemic.

### 3.1 COVimpact interface

The software contains a setting panel, which is divided into two sub-panels. The first subpanel allows the user to choose the French geographical scale (department, region, country) to use for the inputs and outputs. The user can also design the level of protection conferred by natural immunity against severe cases. The motivation for this is that some variants, especially *β* and *γ* could be evading natural immunity [3].

The second subpanel revolves around the vaccination settings. The percentage of the population vaccinated is set separately for three age groups: children from 12 to 17 years old, adults from 18 to 65, and adults above 65. These classes were chosen to reflect differences in infection fatality ratio (IFR, [8]) but also implementation of vaccination strategies. The vaccine efficacy, i.e. the percentage of decrease in severe infections amongst vaccinated people, is set manually and is assumed not to vary with age.

COVimpact allows the user to implement several vaccines in the population. Each vaccine has its own efficacy and its own coverage in the three age classes. Note that individuals are assumed to be vaccinated with only one kind of vaccine, i.e. the sum of the percentage of vaccinated individuals in an age class cannot be greater than 100%.

COVimpact displays 5 types of outputs following the settings provided by the user and an estimation of the proportion of the population already infected, i.e. with natural immunity (see the Methods). The interface is shown in Figure 2. The number of ICU admissions and deaths associated with the epidemic wave (shown in the top panel as a function of the epidemic wave size) are stratified by age in the middle panels. The bottom panel shows the age-stratification of the inferred prevalence of natural immunity. In all the figures, the confidence intervals shown are solely derived from the confidence intervals associated with the estimation of the percentage of the population already infected.

**Fig 1:**
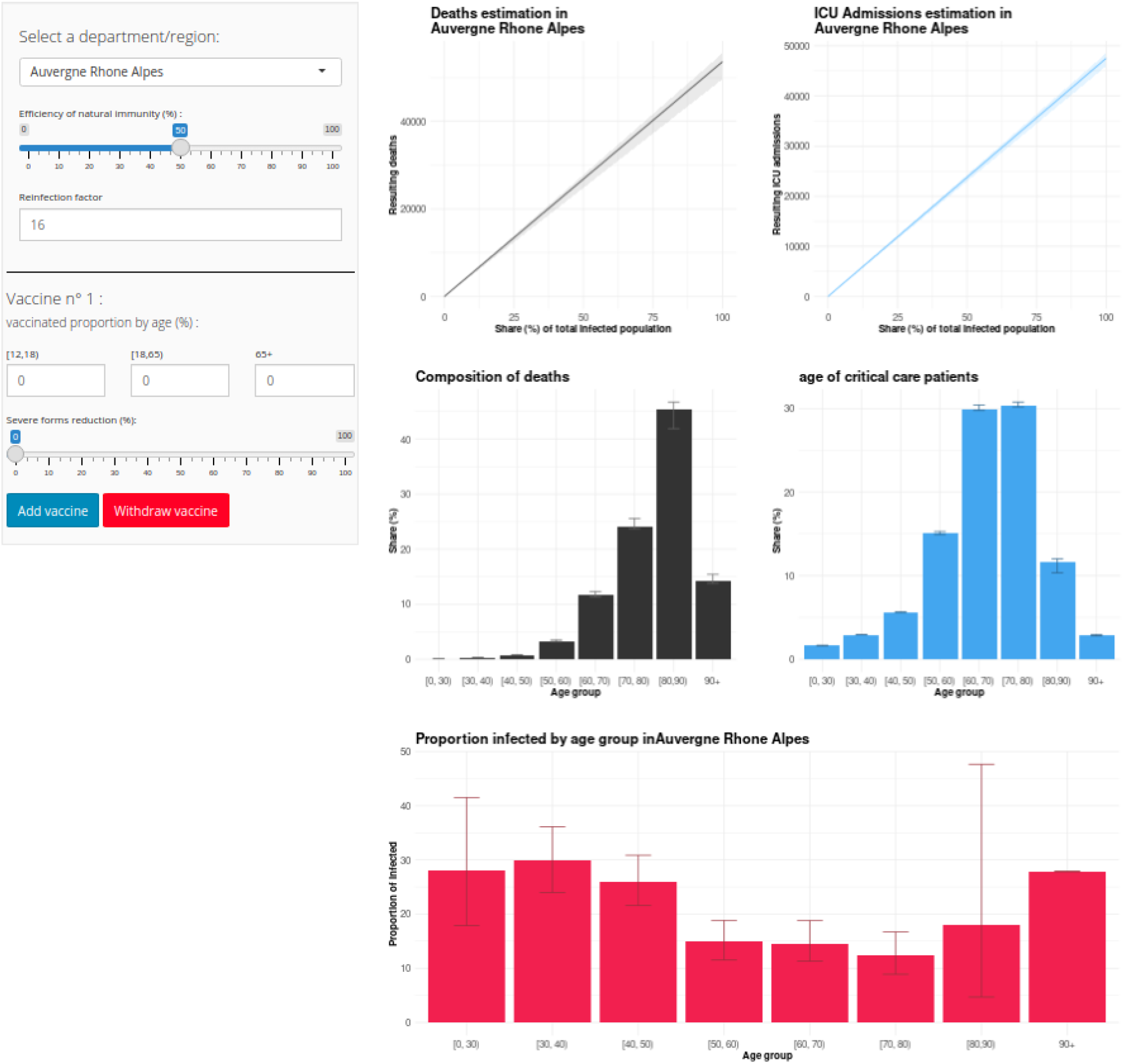
COVimpact user window.

**Fig 2:**
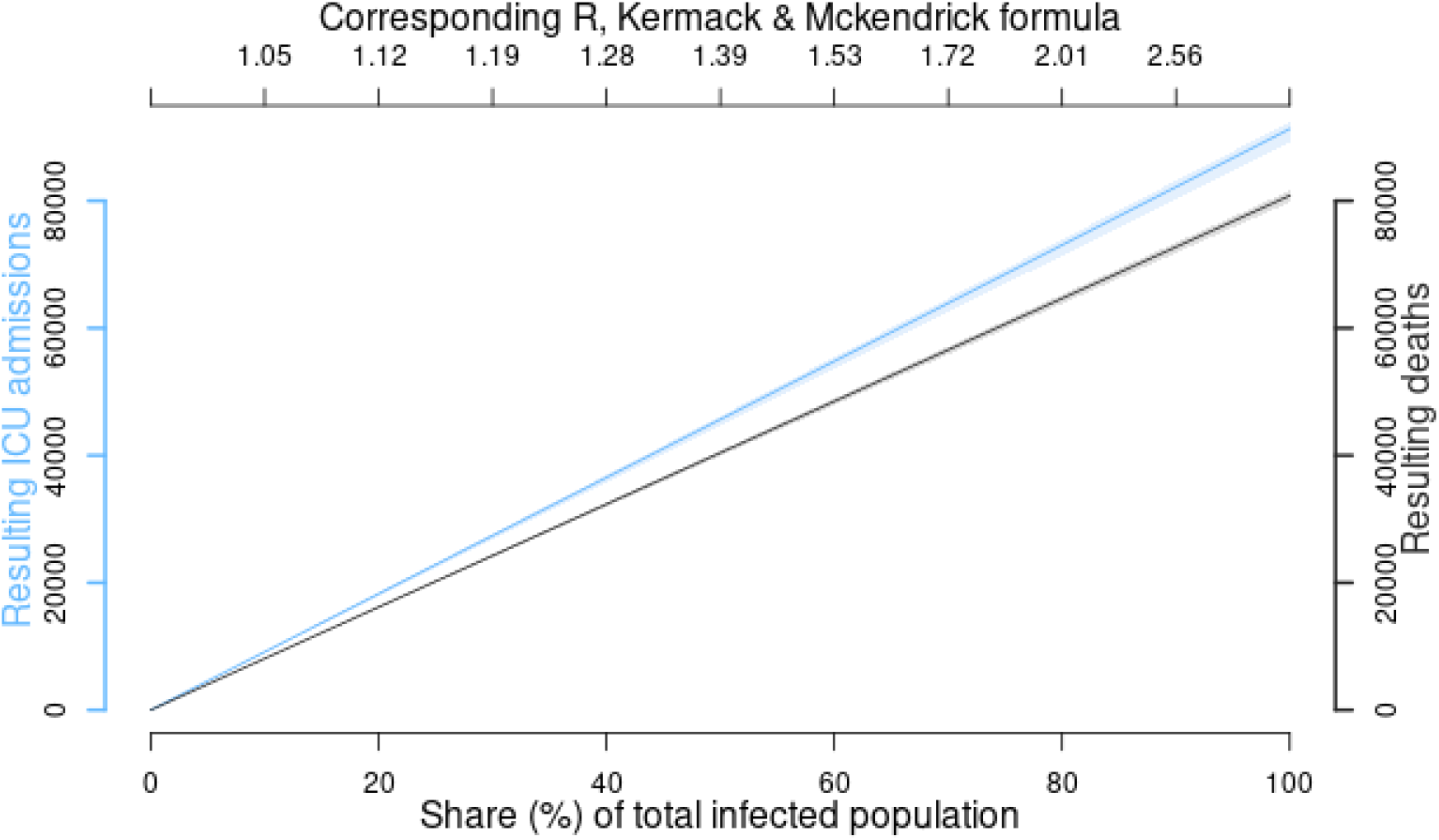
Total number of ICU admissions (in blue) and death (in black) as a function of epidemic wave size in France in 2021. For this figure, we assume a 95% vaccine coverage in adults with an 85% efficiency against severe forms. On the top x-axis, using equation 1, we show the reproduction number associated with the amplitude of the epidemic wave. Confidence intervals on the figure originate from the uncertainty in the estimation of the number of hosts with natural immunity.

### 3.2 France as a test case

To illustrate the implications of the insights provided by COVimpact, we investigate the number of ICU admissions that an epidemic wave affecting 25% of the population would cause in France. The associated reproduction number is 1.15, which is consistent with observed dynamics in France [9] and the fact that vaccination only partly limits reinfection and transmission from reinfected hosts [7].

Even with 95% of the adults have a full vaccination scheme leading to an 85% protection against severe symptoms of both vaccine and natural immunity [7], such a wave would cause approximately 20,000 ICU admissions. Since the median duration of a stay in ICU is 14 days, this corresponds to 280,000 days of ICU bed occupancy. Even if the epidemic wave lasted 2 months, this would mean more than 4,600 beds dedicated to COVID-19 patients, i.e. more than half of the ICU capacity, with a peak after a month. Unfortunately, vaccination coverage is currently much lower than 95% in France.

## 4 Discussion

Forecasting epidemic trends more than 4 weeks in advance is essential, e.g. to hire or train qualified personnel. Unfortunately, variations in public health policies make mathematical models unreliable to anticipate long-term trends [9]. We show that valuable insights can be gained in absence of a detailed description of the epidemic dynamics. More precisely, assuming that the size of the epidemic wave and that the vaccine coverage and efficiency are known, it is possible to estimate the total number of ICU admissions and deaths caused by the wave.

The semi-quantitative results we obtain are particularly useful for public health agencies. Indeed, they allow one to explore extreme scenarios to make informed decisions. For instance, we show that even with extremely high vaccination coverage, and although vaccination strongly decreases the pressure on the health care system, a country like France is likely to experience major strain in ICU occupancy in the event of a large epidemic wave (25% of the population infected in 2 months). To maximise the popularisation of this approach, we also introduce COVIDici, a software allowing one to visualise the effects of an epidemic wave at the French departmental, regional, and national level.

Importantly, our approach makes a series of simplifying assumptions that need to be carefully weighed. First, individuals are assumed to be vaccinated independently of their natural immunisation status. Furthermore, the infection fatality ratio is kept constant as in [8] but this is known to depend on hospital capacity strain [6] or the infecting variant [1]. Second, there were also some missing data at the departmental level, e.g. the age-stratified proportion of infected individuals. These were obtained by adjusting the regional data to the departmental population. Third, regarding the type of immunity, we assume that the protection, quantified as the reduction of severe disease probability, in a vaccinated individual with natural immunity was equal to the rate of the most effective protection. Finally, infected individuals are assumed to be randomly selected in the population, regardless of their level of protection. In other words, the protections (conferred by a past infection, a vaccine or both) only offer a reduction of severe forms but do not affect the risk of being exposed. There are several ways in which this work could be extended. We focused here on France but these insights could be equally valuable for other countries. The main difficulty, which we solved by leaning on earlier work by Hozé et al. [4], is to estimate the proportion of individuals already infected per age class before the epidemic wave. However, the importance of this assumption decreases as the protection conferred by natural immunity decreases. Another extension, more specific to our software, would be to improve the calculations to implement the method by Hozé et al. [4] to re-estimate these proportions but, for France, this requires access to restricted data.

Overall, analysing the consequence of epidemic waves of given sizes as a function of vaccination coverage can greatly help shape public health policies, e.g. to optimise the number of beds in ICU in a region or to estimate the need to hire or train qualified personnel.

## Data Availability

The data used in the study is publically available and the scripts used will be posted on a GitLab server.

https://cloudapps.france-bioinformatique.fr/covimpact/

## Acknowledgements

We thank the ETE team for discussion and the Institut Français de Bioinformatique (IFB) for hosting the software. This work was supported by the PhyEpi grant from the Région Occitanie and the ANR.

