## Supplementary Methods for "Anticipating the hospital burden of future COVID-19 epidemic waves"

### S1 Supplementary Methods

#### S1.1 Calculating the total number of individuals already infected (past attack rate)

For the selected area (department, region, or country), we first initialise the population by removing the individuals in aged care facilities (i.e. in France known as EHPADs), stratifying by age, and estimating, for each age class, the number of people already infected. This latter step relies on the methods implemented by Hozé et al. [3].

More precisely, we extrapolate our inference approach implemented for our COVIDici software [1] to the age-stratified total number of infections estimated by Hozé et al. [3] on April 15, 2021. We calculate the proportion $c$ of the susceptible population that has been contaminated between April 15 and today using the time series of contamination provided by COVIDici.

Mathematically, we have

$$c = \frac{I_{t=0} - I_{t=15/04}}{\text{Pop} - I_{t=15/04}} \tag{S1}$$

where $t = 0$ is the time of interest (after April 15), $I_t$ s the total number of individuals infected at time $t$, and $Pop$ is the total population size. Table S1 shows all the notations used.

From this, we can obtain $I_a$ the total number of individuals of age $a$ infected at the time of interest ($t = 0$, which is dropped from the notations unless stated otherwise for clarity). As indicated above, this is inferred from the accurate estimate from Hozé et al. [3], which is

Table S1: **List and description of the main variables used.** A reference is added for key estimates taken from the literature.

| Variable | Meaning |
|---|---|
| $t = 0$ | Current date |
| $t =$ "15/04" | April 15 2021 |
| $c$ | Correction factor to adjust April 2021's contamination estimation to the current date |
| Pop | target territory's population size |
| $X_a$ | Variable $X$ for age group $a$ |
| $I^*$ | Number of infections estimated on April 15, 2021, by Hozé et al. [3] |
| $I$ | Number of infections for age group $a$ at date $t$ |
| $M$ | Proportion of individuals with post-infection immunity (16% reinfection [2]) |
| $N$ | Number of different vaccines (user input) |
| $A$ | Number of age groups |
| $W$ | Size of a protection-free population equivalent to the protected population |
| $\mathbb{NI}$ | Individuals with natural immunity |
| $\mathbb{R}$ | Individuals with only natural immunity (no vaccine) |
| $\mathbb{V}_i$ | Individuals vaccinated with vaccine $i$ |
| $\mathbb{Q}_i$ | Individuals with only vaccine $i$ as protection (no previous infection) |
| $\mathbb{S}$ | Individuals without protection (neither a vaccine, nor previous infection). |
| $\text{freq}(\mathbb{X})$ | frequency of host category $\mathbb{X}$ with respect to the whole population |
| $\mathscr{E}(\mathbb{X})$ | protection efficiency for category $\mathbb{X}$ (in terms of severe infection probability reduction) |
| $\text{ICU}_{\text{total}}$ | the total number of ICU admissions expected if the entire population was infected |
| $P[\text{ICU}]$ | probability for an infected host to be admitted to an ICU [4] |
| $P[H]$ | probability for an infected individual to be hospitalized [4] |
| $P[D]$ | probability for an individual to die from the infection [4] |
| $D_{\text{total}}$ | total number of death expected if the entire population was infected |
| $\text{freq}_a(D)$ | proportion of the deaths from all age groups that belong to age group $a$ |

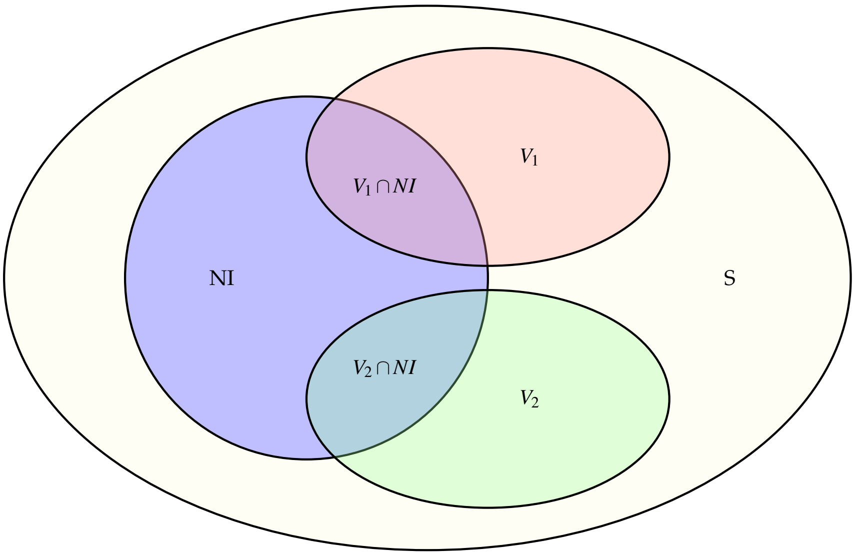

Fig. S1: **Venn diagram of the general structure of the host population.** **S** shows people who are not vaccinated and have not been infected, **NI** are the people with natural immunity due to past COVID19 infection, $\mathbf{V_i}$ are the people vaccinated with the vaccine *i* and $\mathbf{V_i} \cap \mathbf{NI}$ are people vaccinated with the vaccine *i* and have natural immunity due to COVID19 infection in the past.

denoted $I_a^*$. Mathematically, we have:

$$I_a = I_a^* + c\left(\mathrm{Pop}_a - I_a^*\right) \tag{S2}$$

Note that we implicitly assume that from April 15, 2021, to the date of interest, the increase in natural immunity has been the same in proportion for all age classes.

From this, we can calculate the proportion of individuals of age *a* with natural immunity. Importantly, we need to account for the fact that some people (16% according to [2]) are infected more than once, which means the proportion of of individuals with natural immunity is:

$$M_a = 0.84 \frac{I_a}{\mathrm{Pop}_a} \tag{S3}$$

#### Structuring the population into categories

We divide the general population into 6 categories depending on their infection history and vaccination statuses (Figure S1. We then calculate the frequencies of these categories for each age group.

If we denote by $\mathrm{freq}(\mathbb{X})$ the frequency of category $\mathbb{X}$ in the population, we have:

$$\mathrm{freq}(\mathbb{R}_a) = \mathrm{freq}(\mathbb{NI}) - \sum_{i=1}^{N} \mathrm{freq}(\mathbb{V}_{i,a} \cap \mathbb{NI}_a) \tag{S4}$$

$$\mathrm{freq}(\mathbb{Q}_{i,a}) = \mathrm{freq}(\mathbb{V}_{i,a}) - \mathrm{freq}(\mathbb{V}_{i,a} \cap \mathbb{NI}_a) \tag{S5}$$

$$\mathrm{freq}(\mathbb{S}_a) = 1 - \left( \mathrm{freq}(\mathbb{R}_a) + \sum_{i=1}^{N} \mathrm{freq}(\mathbb{V}_{i,a}) \right) \tag{S6}$$

$$\mathscr{E}(\mathbb{V}_i \cap \mathbb{NI}) = \max\left(\mathscr{E}(\mathbb{NI}), \mathscr{E}(\mathbb{V}_i)\right) \tag{S7}$$

where $\mathscr{E}(\mathbb{X})$ is the reduction in severe forms of the infection for category $\mathbb{X}$. By definition, susceptible hosts without a history of infection are unprotected (i.e. $\mathscr{E}(\mathbb{S}) = 1$). Note that in equation S7 we assume that for individuals with both natural and vaccine immunity, the resulting efficiency is that of the most protective between the two.

For each category, we use $\mathscr{E}$) to calculate the (theoretical) number of unprotected hosts $\mathbb{S}$ the category size would correspond to. For example, 10 individuals with 90% protection correspond to one fully unprotected individual.

Then, for a given age class $a$, we compute the total number of unprotected individuals in all the categories, which we denote $W_a$.

$$W_a = \text{Card}(\mathbb{S}_a) + \text{Card}(\mathbb{R}_a)(1-\mathscr{E}(\mathbb{R}_a)) + \sum_{i=1}^{N} \text{Card}(\mathbb{V}_{i,a} \cap \mathbb{NI}_a)(1-\mathscr{E}(\mathbb{V}_{i,a} \cap \mathbb{NI}_a)) + \sum_{i=1}^{N} \text{Card}(\mathbb{Q}_{i,a})(1-\mathscr{E}(\mathbb{Q}_i)) \quad \text{(S8)}$$

where $\text{Card}(\mathbb{X})$ is the cardinal of the set $\mathbb{X}$, i.e. the population size of the category. Note that in equation S8 we assume that vaccine efficacy is the same across all ages (i.e. $\mathscr{E}(\mathbb{Q}_{i,a}) = \mathscr{E}(\mathbb{Q}_i)$).

Finally, we apply the age-stratified probabilities of ICU admission and death following SARS-CoV-2 infection to these population using estimates from Salje et al. [4]. This allows us to compute the total number of ICU admissions ($\text{ICU}_{\text{total}}$) and deaths ($D_{\text{total}}$) caused by the epidemic wave. We also stratify these numbers by host age. Mathematically, we have:

$$\text{ICU}_{\text{total}} = \sum_{a=1}^{A} P[\text{ICU}|H]_a P[H]_a W_a \quad \text{(S9)}$$

$$D_{\text{total}} = \sum_{a=1}^{A} P[D]_a W_a \quad \text{(S10)}$$

$$\text{freq}_a(D) = \frac{P[D]_a W_a}{D_{\text{total}}} \quad \text{(S11)}$$

$$\text{freq}_a(\text{ICU}) = \frac{P[\text{ICU}|H]_a P[H]_a W_a}{\text{ICU}_{\text{total}}} \quad \text{(S12)}$$

All these calculations are performed as soon as the input parameters are modified in the input panel.

where the $P$ stand for probabilities to be hospitalized ($H$), admitted to ICU, or die ($D$), as detailed in Table S1.
